# Privacy-Preserving Matching for Federated Causal Inference in Multicentre Patient Cohorts

**DOI:** 10.64898/2026.07.16.26358171

**Authors:** Roy Gusinow, Andrei Scott Morgan, Lorenzo Maria Canziani, Jennifer Zeitlin, Mihyeon Kim, Elisa Gentilotti, Jade Ghosn, Aline-Marie Florence, Adriana Tami, Alice Toschi, Zaira R. Palacios-Baena, Evelina Tacconelli, Jan Hasenauer

**Affiliations:** The Life and Medical Sciences Institute (LIMES), University of Bonn, Germany; Bonn Center for Mathematical Life Sciences, University of Bonn, Germany; Obstetric, Perinatal, Paediatric and Lifecourse Epidemiology (OPPaLE) team, Centre for Research in Epidemiology and Statistics (CRESS) UMR1153, INSERM, Paris, France; Elizabeth Garrett Anderson Institute for Women’s Health, UCL, London, UK; Division of Infectious Diseases, Department of Diagnostics and Public Health, University of Verona, Verona, Italy; Université Paris Cité and Université Sorbonne Paris Nord, INSERM UMR 1137 IAME, Paris, France; Infectious and Tropical Diseases Department, Bichat – Claude Bernard Hospital, Assistance Publique - Hôpitaux de Paris, Paris, France; Department of Epidemiology, Biostatistics and Clinical Research, Bichat – Claude Bernard Hospital, Assistance Publique-Hôpitaux de Paris, Paris, France; University of Groningen, University Medical Center Groningen, Department of Medical Microbiology and Infection Prevention, Groningen, The Netherlands; Infectious Diseases Unit, IRCCS Azienda Ospedaliero-Universitaria di Bologna, Bologna, Italy; Department of Medical and Surgical Sciences, University of Bologna, Bologna, Italy; Unidad Clínica de Enfermedades Infecciosas y Microbiología, Hospital Universitario Virgen Macarena; Departamento de Medicina, Universidad de Sevilla; Instituto de Biomedicina de Sevilla (IBiS)/CSIC, Seville, Spain; CIBERINFEC, Instituto de Salud Carlos III, Seville, Spain

**Keywords:** Federated Learning, Differential Privacy, Causal Effects, Matching Methods

## Abstract

**Motivation:** Causal effect estimates can often be biased in clinical and epidemiological studies as patient cohorts frequently exhibit substantial covariate imbalances between treated and control groups, often amplified in multicentre studies due to heterogeneous recruitment, clinical practice, and case mix. Covariate balancing methods are therefore essential for valid causal inference. However, their application becomes challenging when data are distributed across cohorts and cannot be pooled because of privacy, legal, or institutional constraints, leaving a gap in practical methods for causal effect estimation in federated and imbalanced clinical data settings.

**Results:** We develop a privacy-preserving framework for covariate balancing and causal effect estimation across distributed data providers, combining federated aggregation with differential privacy to enable propensity score subclassification and matching without sharing individual-level records. Matching relies on non-disclosive quantities and differentially private distance evaluation, and the resulting matched subsets remain local to each server. Balance can be assessed through federated diagnostics and privacy-preserving visualisations, and we provide secure estimators for average treatment effects with associated uncertainty quantification. We implement this framework in the DataSHIELD federated analysis platform via 2 R packages. In simulations, we demonstrate agreement between federated and centralised analyses in the absence of privacy noise and quantify the bias–variance trade-offs induced by differential privacy. We illustrate applicability in two multinational settings—a Long COVID cohort and very preterm birth cohorts—showing that the approach enables practical causal analyses under real-world data protection constraints.

**Availability and Implementation:** The DataSHIELD packages are available on Github (https://github.com/roygusinow/dsMatchingClient, https://github.com/roygusinow/dsMatching). Additional methodological details are provided in the Supplementary Material.

**Contact:** Jan Hasenauer

## Introduction

Observational studies play a central role in assessing the effects of exposures and interventions in clinical and epidemiological research. Unlike randomised trials, however, treatment and control groups in observational cohorts often differ systematically in baseline characteristics due to non-random sampling, clinical decision-making, or structural differences in recruitment. Such covariate imbalances can induce confounding and lead to biased estimates of causal effects if not appropriately addressed. This challenge persists even in large-scale multicentre and multinational consortia, where increased sample size is frequently accompanied by substantial heterogeneity in patient populations and study protocols.

Matching and related covariate balancing methods are widely used to mitigate confounding by constructing treated and control groups with similar distributions of observed characteristics. These methods aim to approximate the conditions of a randomised experiment, thereby enabling more interpretable causal effect estimation. Matching has become a standard tool across disciplines, including econometrics [Rosenbaum and Rubin, 1983], epidemiology [Brookhart et al., 2006], political science [Ho et al., 2007], sociology [Morgan and Harding, 2006], and psychology [Steiner and Cook, 2013]. When successful, matching reduces reliance on model extrapolation and provides a transparent diagnostic framework for assessing covariate balance prior to downstream analyses.

In practice, however, the application of matching methods in clinical research is increasingly constrained by data governance considerations. Patient-level data are often collected and stored across multiple sites, such as hospitals or national registries, and established legal and ethical frameworks limit direct data sharing [Hansen et al., 2021]. This can substantially slow analyses, reduce effective sample size, and limit statistical power, with potentially harmful consequences for clinical decision-making and patient outcomes [Bentzen et al., 2021]. As a result, there is a growing need for analytical methods that support robust causal inference while respecting data sovereignty and privacy requirements. Conceptually, there are two different frameworks: meta-analysis and federated analysis. While meta-analysis methods have been successfully used to combine results from independent studies, correct aggregation of values to estimate treatment effects requires considerations of the underlying heterogeneous distributions and large sample sizes such that point estimations and confidence intervals are not biased [Higgins et al., 2009, Rice et al., 2018]. In addition, such meta-analyses do not balance populations across sites or studies, which may be particular relevant when treated and control individuals are unevenly distributed, since comparable controls for one site may be located in another site and thus would be inaccessible without privacy safeguards.

One approach to addressing these constraints is privacy-preserving federated analysis, in which statistical models are fitted across distributed datasets without centralising individual-level data. Federated learning enables the estimation of global model parameters by exchanging only non-disclosive, aggregated information between local data holders and a central coordinator, often through iterative optimisation procedures such as gradient averaging [McMahan et al., 2017]. Complementary privacy guarantees can be provided through differential privacy, whereby carefully calibrated noise is added to communicated quantities to limit the risk of information leakage [Dwork, 2006]. Together, these approaches allow collaborative analyses across institutions while maintaining formal privacy protections. There is a growing ecosystem of federated methods, ranging from tailored implementations for specific application domains (e.g., medical image analysis) to general-purpose platforms for statistical and machine-learning workflows, including DataSHIELD [Avraam et al., 2025], TensorFlow Federated [Abadi et al., 2015], and Swarm Learning [Warnat-Herresthal et al., 2021].

In this work, we introduce a collection of privacy-preserving matching methods for covariate balancing across distributed datasets. The proposed methods are implemented in DataSHIELD [Avraam et al., 2025] through two new packages, *dsMatching* and *dsMatchingClient*. These packages combine federated learning and differential privacy techniques to provide secure implementations of commonly used matching workflows, with functionality analogous to established tools for centralised data analysis, e.g. *MatchIt* [Ho et al., 2011] and *marginaleffects* [Arel-Bundock et al., 2024]. These include custom matching specifications, non-disclosive pre- and post-match statistics, plotting functionalities for visualising covariate balance, as well as estimating average treatment effects and associated standard errors.

We evaluate the proposed methods in a simulation study to quantify the impact of differential privacy on matching performance and treatment effect estimation, and demonstrate their practical utility in two real-world applications: the multinational ORCHESTRA Long COVID cohort [Gentilotti et al., 2023] and three population-based very preterm birth cohorts [Larroque et al., 2004, Ancel and Goffinet, 2014, Zeitlin et al., 2020] accessed via the RECAP Preterm platform[Bamber et al., 2022].

## Methods

We consider the problem of estimating the causal effects of a binary exposure or intervention from observational studies. Therefore, let *D* ∈ {0, 1} denote a treatment indicator, *X* a vector of observed baseline covariates, and *Y* an outcome of interest. Throughout, we assume that causal effects are identified conditional on *X*, and focus on methods that address systematic differences in the distribution of *X* between treated and untreated groups. The proposed methods should be interpreted as addressing measured confounding only.

### Centralised covariate balancing and causal effect estimation

In observational data, treated and untreated individuals often differ systematically in baseline characteristics, leading to covariate imbalance and confounding. If not appropriately addressed, such imbalances can induce biased estimates of causal effects. Covariate balancing methods aim to mitigate this problem by constructing treated and control groups with similar distributions of observed covariates.

A widely used approach for covariate balancing is propensity score–based adjustment. The propensity score

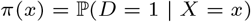

encapsulates the probability of treatment assignment conditional on observed covariates. The estimation of a sample’s propensity can be formulated as a logistic regression exercise, where the confounders *X* are regressed on the treatment indicator *D*. Under standard assumptions, conditioning on the propensity score balances the distribution of *X* between treatment groups.

Common strategies for using propensity scores include:

(i) subclassification, in which individuals are grouped into strata defined by quantiles of the estimated propensity score, and (ii) matching, in which treated and untreated individuals with similar propensity scores are paired or grouped to form a balanced subset of the data. The resulting matched data are then typically investigated via summary statistics tables and visualisation aids to assess the balance of covariates. After this has been achieved, causal estimands such as the average treatment effect (ATE), the average treatment effect on the treated (ATT), or the average treatment effect on the untreated (ATU) can be estimated using outcome models or weighted contrasts.

### Consideration for a transition from centralised to federated analysis

To facilitate causal effect analyses for distributed clinical and epidemiological datasets, we consider a federated setting in which individual-level data remain at their hosting sites and only aggregated, non-disclosive information can be exchanged. In this setting, key steps of the centralised workflow— including propensity score estimation, covariate balancing via subclassification or matching, and downstream effect estimation—must be redesigned to avoid sharing individual-level covariates, outcomes, or cross-site pairwise comparisons. Moreover, repeated communication of intermediate quantities can create disclosure risks, requiring explicit privacy safeguards. These safeguards include preventative action when the number of samples used in aggregation are below the threshold set by the data custodian, which a standard procedure set for most DataSHIELD functionalities (Supplementary Note 2.1).

We consider the primary adversary to be an authenticated but malicious analyst operating through the central client. This adversary can issue permitted DataSHIELD commands, observe all quantities returned to the client, and combine outputs across repeated calls. Their goal is to infer individual-level information stored at remote sites, including exact covariate values, treatment or outcome status. We assume that remote data sites do not intentionally disclose individual-level records, and that network communication between client and server is protected by the secure platform infrastructure. The privacy safeguards introduced below are therefore designed to reduce disclosure through client-visible outputs under authenticated but potentially adversarial use of the analysis interface.

Federated implementations must therefore satisfy the following requirements: (i) local computation of individual-level quantities such as propensity scores; (ii) aggregation-only communication, where only summary statistics or perturbed quantities are transmitted, such as those which compose the covariate balance diagnostics and plotting functionalities; (iii) protection against linkage and reconstruction attacks, for example through randomisation, shuffling, or differential privacy mechanisms; and (iv) consistency with centralised estimands, ensuring that the federated workflow targets well-defined causal quantities.

Finally, the current workflow assumes that the variables required for propensity score estimation, matching, and outcome modelling are available as complete cases. This follows from the federated GLM implementation used for propensity score and outcome model estimation, which excludes observations with missing treatment, outcome, or covariate values. Consequently, as in centralised complete-case propensity score analyses, estimates may be biased when missingness is informative or differs across sites. In practice, missingness rates should be assessed before matching using non-disclosive site-level summaries. Federated imputation methods, including multiple imputation by chained equations as implemented in the base DataSHIELD package, may also be applied before matching, with the matching and effect-estimation workflow recommended to be repeated across imputed datasets [Azur et al., 2011, Avraam et al., 2025].

### Privacy-preserving federated subclassification and matching

To enable covariate balancing across distributed datasets under privacy constraints, we developed two complementary approaches: federated subclassification and federated matching. Both approaches retain individual-level data at each site and rely on exchanging only non-disclosive information.

**Federated subclassification** extends standard (centralised) propensity score subclassification to the distributed setting in a straightforward manner. We define strata using quantiles of the global propensity score distribution, estimate the corresponding quantile boundaries in a federated manner, and communicate these boundaries to all sites. Each site assigns its local individuals to subclasses based on the shared boundaries and communicates stratification-based weighting based solely on subclass sizes, thereby achieving covariate balancing without sharing individual-level covariates, outcomes, or propensity scores. The number of subclasses is specified by the analyst, allowing users to evaluate multiple balancing outcomes. As with most functionalities, the number of samples within each subclass must reach the minimum threshold set by the data custodian in order to proceed with the action. Thus, the quantiles specified are restricted depending on the distribution of samples within each remote server.

**Federated matching** approximates standard propensity score matching while avoiding cross-site sharing of individual-level records. Since matching requires comparing propensity scores across sites, we introduced a privacy-preserving two-stage procedure. First, each site estimates local propensity scores and releases a differentially private version,

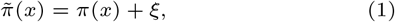

where *ξ* ~ Lap(0, *b*) is the Laplace distribution with scale *b* = 1*/ϵ* where *ϵ* denotes a site-defined privacy budget (Figure 1a). Second, sites transmit only the perturbed propensity scores, together with randomised and shuffled identifiers and treatment indicators, to a coordinating entity that performs the matching procedure using 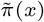 as the distance measure (Figure 1b). The coordinating entity returns the identifiers of the selected matches, and each site constructs the matched subset locally (Figure 1c).

**Fig. 1.**
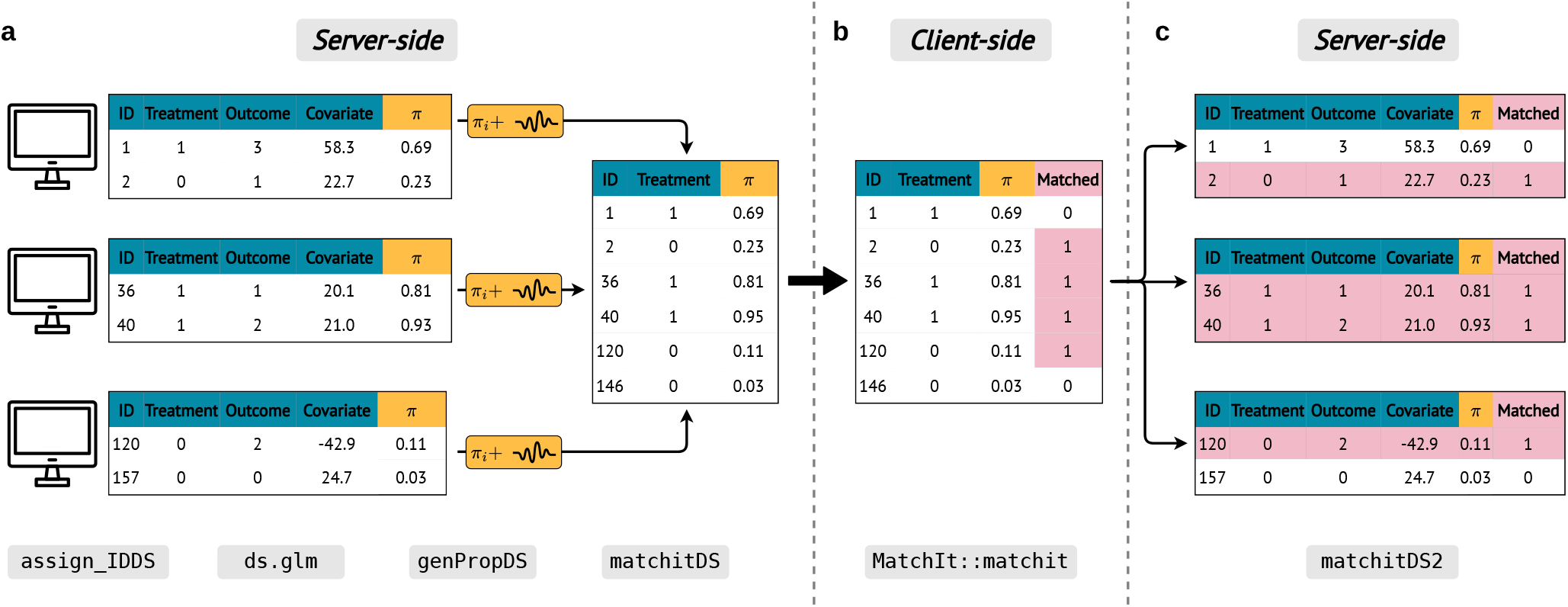
Illustration of the Functionality of Federated Matching. (a) A randomly-generated ID is generated for each sample in all nodes at the server-side, and propensity scores are generated for individual samples. The differentially private propensity scores are communicated to the central client. (b) At the client-side, the IDs, treatment and propensity scores are pooled and then utilised by the matching algorithm. (c) Samples selected after matching then relay this assignment to each server for use in post-matching evaluation metrics.

The federated generalised linear model (GLM) implementation from the base DataSHIELD package utilises aggregated gradients of the likelihood from each remote site, requiring several iterative communication rounds. These quantities are returned as aggregated, disclosure-controlled outputs rather than individual-level records, and potentially disclosive quantities such as individual-level residuals or linear predictors are not returned by the standard DataSHIELD implementation [Gaye et al., 2014, Avraam et al., 2025]. Thus, the differential privacy mechanism and corresponding budget is specified only for the sending of propensity scores to the central site, as it is not required for sharing gradients and other aggregated quantities such as summary statistics or outcomes (Supplementary Note 2.2).

This design prevents disclosure of individual-level covariates and exact propensity scores, while enabling approximate matching across distributed cohorts through privacy-preserving communication of perturbed distances.

### Federated estimation of causal effects

Building on federated subclassification and federated matching, we estimate causal effects on covariate-balanced populations while keeping all individual-level data at their hosting sites. Subclass assignments, weights, and matched-set membership are constructed locally, and causal effect estimation relies on aggregating only non-disclosive quantities across sites.

To estimate causal effects in a federated manner, we adopt a *g-computation* framework [Chatton et al., 2020] in which the outcome *Y* is modelled as a function of the treatment indicator *D* and covariates *X*. Similar to propensity score estimation, the parameters of the outcome model *Y* | *D, X* are estimated using federated optimisation, where gradients are computed locally at each site and aggregated across sites during each optimisation step [Gaye et al., 2014, McMahan et al., 2017]. This procedure yields global parameter estimates 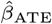 without sharing individual-level outcomes or covariates.

A commonly used causal estimand is the average treatment effect (ATE), defined as

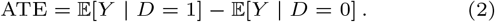

We estimate the ATE using marginal mean weighting through propensity score stratification (MMWS) [Hong, 2010]. Specifically, treatment effects are first estimated within each propensity score subclass, and subclass-specific expectations are then weighted by the proportion of individuals in the corresponding treatment group within each stratum. These weighted estimates are computed locally and aggregated across sites according to the relative subclass composition, yielding a global ATE without centralising sensitive data.

The same framework naturally extends to related causal estimands, including the average treatment effect on the treated (ATT), the average treatment effect on the untreated (ATU), as well as risk ratios (RR) and odds ratios (OR), by restricting to the relevant subpopulations and applying the appropriate contrast functions to the estimated mean potential outcomes (Supplementary Note 2.3).

We note that g-computation and MMWS in a federated setting introduces additional computational and coordination complexity compared to centralised analyses. Outcome models, subclass-specific effects, and weighting schemes must be evaluated locally, aggregated across heterogeneous site sizes, and communicated in a non-disclosive manner, while preserving consistency with the corresponding centralised estimands.

### Cluster-robust standard errors of treatment effect estimands

To quantify uncertainty in treatment effect estimands, we account for the heteroskedastic nature of observations within the experiment, as well as the dependency of observations after subselection. Following established recommendations (for centralized computations), we compute cluster-robust standard errors [Abadie and Spiess, 2022].

We employ the delta method and use a first-order Taylor approximation of the treatment effect estimator (Equation 2) to propagate uncertainty from estimated model parameters to the causal estimands [Casella and Berger, 2024]. In the federated setting, sites compute the required aggregated components locally and communicate only non-disclosive summaries. Specifically, each site contributes aggregated terms for the Jacobian and the cluster-robust variance– covariance matrix, which are then combined to obtain global uncertainty estimates. We then use the sandwich estimator to calculate the global dispersion and estimating function, with options available for heteroskedasticity-consistent standard error types (HC0 and HC1). HC0 corresponds to the unadjusted sandwich variance estimator and is generally appropriate when the number of observations is large, whereas HC1 applies a finite-sample degrees-of-freedom correction and is generally preferable in smaller sample settings. This provides uncertainty quantification comparable to centralised analyses while ensuring that individual-level outcomes, covariates, and cluster-level information remain private.

### Evaluating post-matching covariate balance

To evaluate the effectiveness of federated matching and to assess whether covariate balance has been achieved between treated and control groups, we compute standard balance diagnostics before and after matching. These include weighted means, standardised differences, variance ratios, and empirical cumulative distribution function (eCDF) comparisons between treatment groups [Ho et al., 2011].

In the federated setting, each site computes balance diagnostics locally and communicates only aggregated, non-disclosive summaries, which are then combined to obtain cohort-level assessments. To support interpretation, we construct visual diagnostics from aggregated quantities, including scatter plots of standardised differences across covariates as well as quantile–quantile, density, and eCDF plots. In line with recommended non-disclosive visualisation practices, these plots rely on aggregated summaries and a limited number of evaluation points, ensuring that individual-level data are not leaked while enabling effective evaluation of post-matching covariate balance [Avraam et al., 2021].

## Implementation

DataSHIELD provides a server infrastructure software and series of federated R packages that enable non-disclosive analyses [Avraam et al., 2025]. It supports a secure client– server architecture, disclosure controls, and general-purpose federated computation tools. This includes standard operations such as mean calculation through its default R package, dsBase, and there is extended functionality supplied via a variety of specialised toolsets developed by the community. Yet, there are no tools which facilitate covariate balancing, and corresponding downstream diagnostics and outcome effects.

We implement the proposed balancing framework within the DataSHIELD ecosystem following its client–server architecture: individual-level data remain on secure server nodes, while the analyst interacts through a central client that coordinates computations and aggregates non-disclosive results.

In DataSHIELD, all operations involving sensitive variables are executed locally on the servers, and only aggregated, non-disclosive quantities are returned to the client over encrypted HTTPS connections (Figure 2). Matched-set indicators, subclass assignments, and all outcome and covariate data remain local to each site; the client only stores aggregated summaries and derived diagnostics.

**Fig. 2.**
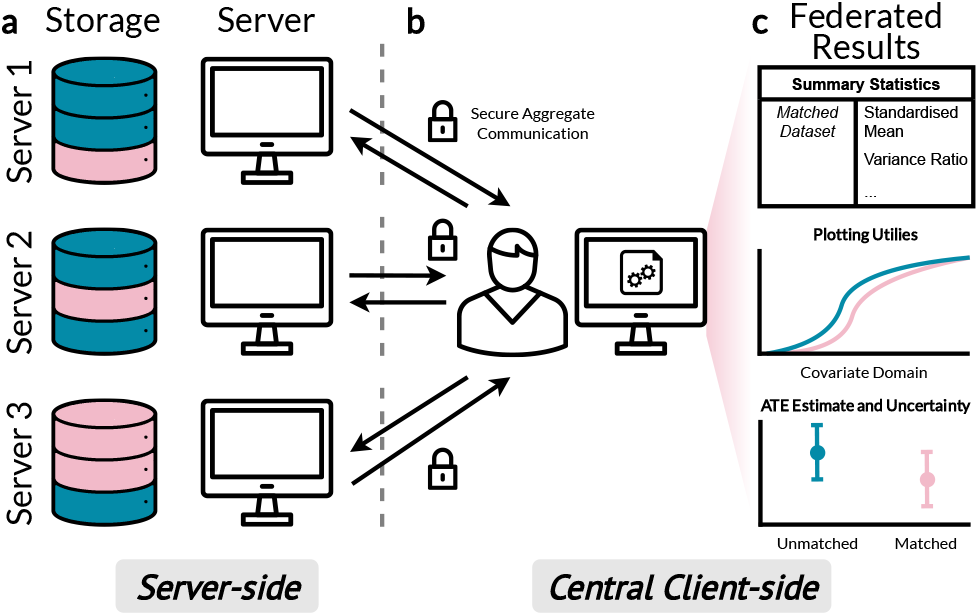
Communication Set-up for Federated Learning Platform. (a) Three servers each contain data of unmatched (blue) and matched (pink) samples, as well as online servers. (b) Only non-disclosive data are sent between the servers and central client via HTTPS. (c) After secure communication with the servers, the central client aggregates the data received. In the framework of *dsMatching*, tables of summary statistics, plotting utilities for visualisation as well as secure treatment effect are provided, similar to the situation when results are computed centrally.

### Core functional components

Our implementation provides four main components that mirror the analytical workflow.

#### Propensity score estimation

Propensity scores are estimated using federated generalised linear models available in the DataSHIELD infrastructure, with model fitting performed via gradient averaging. In this setting, model parameters are estimated through iterative aggregation of site-level likelihood gradients, yielding regression coefficients equivalent to those obtained from a centralised GLM up to numerical precision. Consequently, in the absence of differential privacy noise, the resulting propensity scores are numerically identical to those from the corresponding centralised analysis. Propensity scores are retained locally and used as inputs to balancing procedures.

#### Federated subclassification and federated matching

For federated subclassification, the client aggregates global propensity score quantiles and distributes the corresponding cut-points to sites, which then assign subclass membership locally and compute subclass-specific summaries and weights.

For federated matching, sites compute propensity scores locally and transmit only differentially private, randomised, and shuffled quantities together with non-identifying metadata required for matching. The client performs matching using these privacy-preserving values and returns matched identifiers to each site, where matched subsets are constructed locally.

#### Federated effect estimation

Causal estimands are computed from local subclass- or matched-subset quantities and aggregated at the client. Outcome models are fitted using federated optimisation, and causal contrasts are evaluated using stratification- or matched-set summaries, consistent with the estimands defined in Section 2.4.

#### Diagnostics and uncertainty quantification

Sites compute balance diagnostics and aggregated components required for uncertainty quantification locally. The client combines these summaries to produce cohort-level diagnostics, privacy-preserving visualisations, and cluster-robust standard errors using sandwich estimators and the delta method.

### Software availability

The implementation is provided as two interoperable R packages, separating server-side computation primitives from client-side orchestration and aggregation, dsMatchingClient and dsMatching. The packages are available on GitHub under the MIT license.

## Results

We evaluate the proposed framework using both simulated and real-world data. We first use a controlled simulation study to assess statistical properties and privacy–utility trade-offs. We then apply the methods to two multinational clinical cohorts, illustrating their use in regression-based causal inference for Long COVID and in survival analysis for very preterm birth outcomes.

### Simulation Study verifies Package under Privacy Constraints

To analyse the statistical properties of the proposed federated matching framework and to characterise the privacy–utility trade-offs induced by differential privacy, we first consider a controlled simulation study. Specifically, we assess whether the approach recovers correct causal effects under low privacy distortion, how increasing noise affects matching quality and effect estimation, and whether federated inference agrees with centralised analyses when privacy noise is absent.

We adopt a well-established simulation design commonly used to benchmark matching methods [Austin, 2009], as implemented in the MatchIt package [Ho et al., 2011]. We generate *n* = 1000 observations with nine covariates *X*_1_, …, *X*_9_ ~ *N* (0, 1). Treatment assignment is generated as *D* ~ Bern(*p*) with *p* = *f* (−1.5 + 0.5*X*_1_ + 0.2*X*_2_ + 0.2*X*_4_ − 0.2*X*_5_ − 0.2*X*_7_ − 0.2*X*_8_) where *f* (*·*) denotes the logistic function, inducing confounding between treatment and covariates. Outcomes are generated according to *Y* = −2.8 + *D* + 1.5*X*_1_ + 3*X*_2_ + 5*X*_3_ + 4*X*_4_ + 4*X*_5_ + 4*X*_6_ + *ε* with *ε* ~ *N* (0, 5), yielding a true average treatment effect on the treated (ATT) of 1.

Propensity scores are estimated via logistic regression of *D* on all covariates. To emulate the federated matching setting, we add differential privacy noise to the propensity scores (Equation 1) and perform nearest-neighbour propensity score matching without calipers or discarding. For each value of the Laplace scale parameter *b* ∈ {10^*k*^ | *k* = −5, …, 0}, corresponding to privacy budgets *ϵ* = 1*/b*, we repeat the simulation 1000 times to obtain a distribution of the ATT estimates. This range spans the main operating region of the proposed method, from a high-utility setting in which the perturbed propensity scores closely approximate the unperturbed matching solution, to the more noisy setting in which matching becomes increasingly random and the resulting ATT estimates approach those obtained without effective covariate balancing.

We quantified the impact of differential privacy in the simulation study by comparing the matched set obtained using perturbed propensity scores with the matched set obtained without privacy noise. Using the matched set obtained without noise as a reference, we quantify the proportion of correctly selected samples as *b* increases (decreasing *ϵ*) (Figure 3a). For small values of *b*, matching closely reproduces the reference set, whereas increasing noise leads to progressively more random matching, with the intersection converging to the treated fraction.

**Fig. 3.**
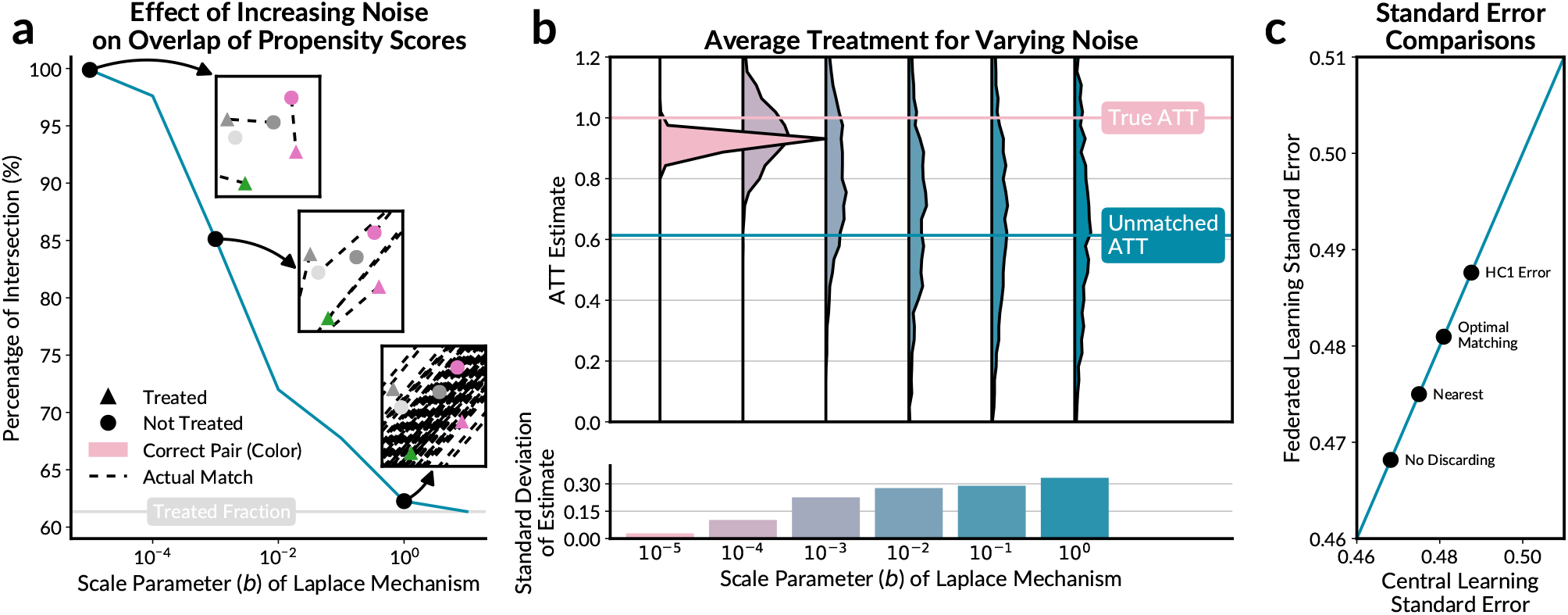
Simulation Study of the Effect of Laplace Scale *b* = 1*/ϵ* on Covariate Balancing. (a) The increase in noise added to the propensity score results in a lower percentage of samples correctly matched. At *b* = 10^0^ (*ϵ* = 10^0^), the percentage intersection is equal to the treated fraction. (b) Effect of scale parameter *b* = 1*/ϵ* on the Average Treatment Effect on the Treated Estimate (ATT). At low noise, the ATT is consistent and close to the true ATT, but as the scale parameter *b* increases, the estimate becomes biased towards lower values and the standard deviation of the ATT estimates across simulations increases. (c) Comparison of central and federated standard estimates across different matching specifications when the privacy budget is infinite (*b* = 0, equivalently *ϵ* = *∞*). Points on the blue line indicated perfect agreement between both approaches.

We next assess treatment effect estimation. Without matching, the naive estimator yields an average ATT of approximately 0.6, which is substantially biased relative to the true effect due to confounding. In contrast, applying the proposed federated matching approach yields ATT estimates close to the true value for small noise levels (Figure 3b). As expected, increasing privacy noise introduces bias and inflates variability, illustrating the privacy–utility trade-off inherent to differentially private matching.

To validate the federated inference machinery, we compare federated and centralised standard error estimates in the absence of privacy noise across several matching specifications, including nearest-neighbour and optimal matching all with HC0 correction, as well as the first heteroskedasticity-consistent variance estimator (HC1). We evaluated whether the federated aggregation of the quantities required for uncertainty estimation reproduces the corresponding centralised calculation when the matched sample is fixed (Figure 3c). Federated and centralised standard errors agree up to machine precision for various matching configurations and outcomes (ATU, ATE, RR, OR) (Supplementary Figure 1), confirming the correctness of the federated implementation.

Overall, the simulation study demonstrates that the proposed framework recovers correct causal effects under low privacy distortion, behaves predictably as privacy constraints increase, and faithfully reproduces centralised inference when privacy noise is absent.

### Non-disclosive Covariate Balancing for Vaccination Efficacy in Post-COVID Patients

To demonstrate the use of the proposed framework for regression-based causal inference in a real-world setting, we analysed the effect of SARS-CoV-2 vaccination on health-related quality of life (HRQoL) among patients with post-COVID condition using data from the ORCHESTRA Long-Term Sequelae Cohort [Gentilotti et al., 2023]. The ORCHESTRA cohort comprises several prospective subcohorts recruited across 56 centres in five European countries, including adults with laboratory-confirmed SARS-CoV-2 infection. Baseline data include demographic characteristics, clinical factors, and treatment-related variables, while follow-up assessments capture persistent symptoms and HRQoL. HRQoL was measured 6 months after infection using the Short Form Health Survey (SF-36), with physical component score being the outcome metric [Hays et al., 1993, Ware Jr, 2000]. Scores below the norm-based reference value (50) indicate impaired quality of life relative to the general population.

For this analysis, we considered five subcohorts hosted at different sites and assumed that individual-level data could not be shared across centres. The subcohorts differ substantially in sample size, vaccination prevalence, and baseline characteristics (Figure 4a), reflecting real-world heterogeneity in recruitment strategies and regional vaccination campaigns. In particular, most vaccinated individuals were enrolled in the UNIVR subcohort, whereas INSERM contributed a large fraction of unvaccinated patients (Figure 4c), leading to pronounced imbalance between treatment groups.

**Fig. 4.**
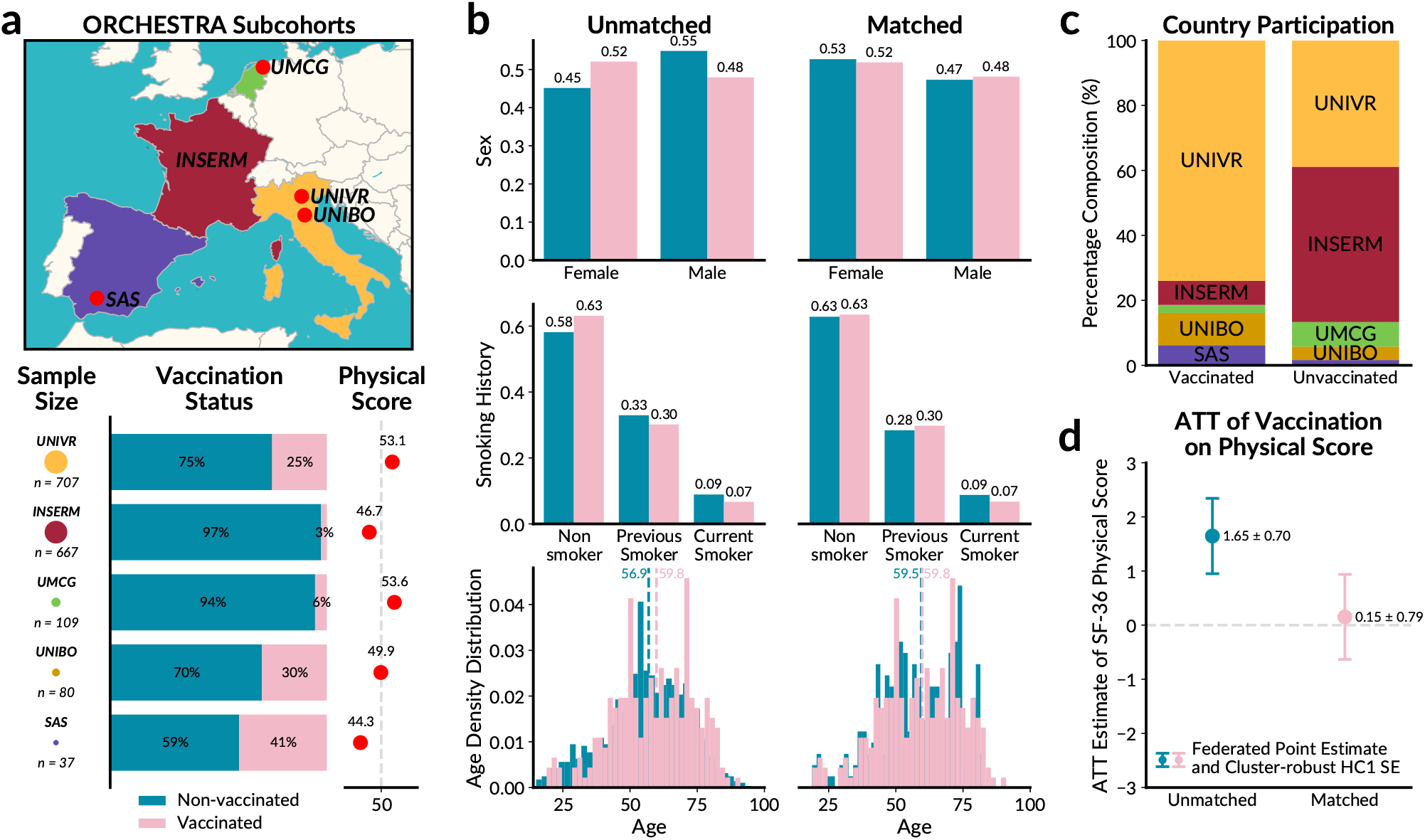
Non-disclosive Covariate Balancing in the Long-Term Sequelae Cohort. (a) Map of the five ORCHESTRA subcohorts as well as their respective sample sizes, percentage frequency of vaccinations and their HRQoL SF36 physical score. (b) Density distributions of demographics variables for non-vaccinated/vaccinated patients before and after the federated matching procedure. Vertical dashed lines in the age distribution indicate respective mean values. (c) Relative contribution of the individual cohorts to the number of non-vaccinated/vaccinated patients. (d) Average treatment effect on the treated and its standard error as calculated in a federated manner for the unmatched and matched sets.

To address this imbalance, we applied federated nearest-neighbour propensity score matching using age, sex, and smoking status as covariates, selected because they were complete across each subcohort.

Federated matching substantially improved covariate balance between vaccinated and unvaccinated patients across subcohorts, aligning demographic distributions without sharing individual-level data (Figure 4b). Before matching, unadjusted analyses suggested a positive association between vaccination and HRQoL.

After federated matching, however, the estimated average treatment effect on the treated was close to zero (Figure 4d), suggesting that the apparent benefit was largely attributable to measured confounding due to cross-cohort imbalance.

This example illustrates how privacy-preserving federated matching enables robust causal inference in heterogeneous, multi-national cohort studies and helps avoid misleading conclusions based on unadjusted comparisons.

Due to the unperturbed propensity scores generated by the analysis not being strongly separable, the resulting matched set and outcome estimate are particularly sensitive to differentially private noise. We set the privacy budget of *ϵ* = 10^5^ for each server, noting that a high budget was chosen to preserve analytical utility as the required privacy permissions for analysing the dataset were already approved. In all applications, the privacy budget should be determined by the relevant data custodians, and analysts should assess whether the selected budget achieves acceptable covariate balance using the federated balance diagnostics provided by the package, similar to the centralised matching case.

### Secure Matching for Evaluating Interventions in Very Preterm Cohorts

As a second application, we investigated the effect of delivery mode (vaginal or Caesarean) on survival to hospital discharge of babies of 500g or more without congenital anomalies born alive between 22 and 31 weeks gestational age (GA). We used three geographically-based preterm birth cohorts included in the RECAP Preterm platform [Bamber et al., 2022]: EPIPAGE1 [Larroque et al., 2004] (France, 1997), EPIPAGE2 [Ancel and Goffinet, 2014] (France, 2011) and EPICE [Zeitlin et al., 2020] (19 regions from 11 European countries, 2011); (Figure 5a). These respective datasets contain differences in the number of samples, delivery modes and survival outcomes (Figure 5b), as well as heterogeneous variables due to the varying locations and time periods of the studies.

**Fig. 5.**
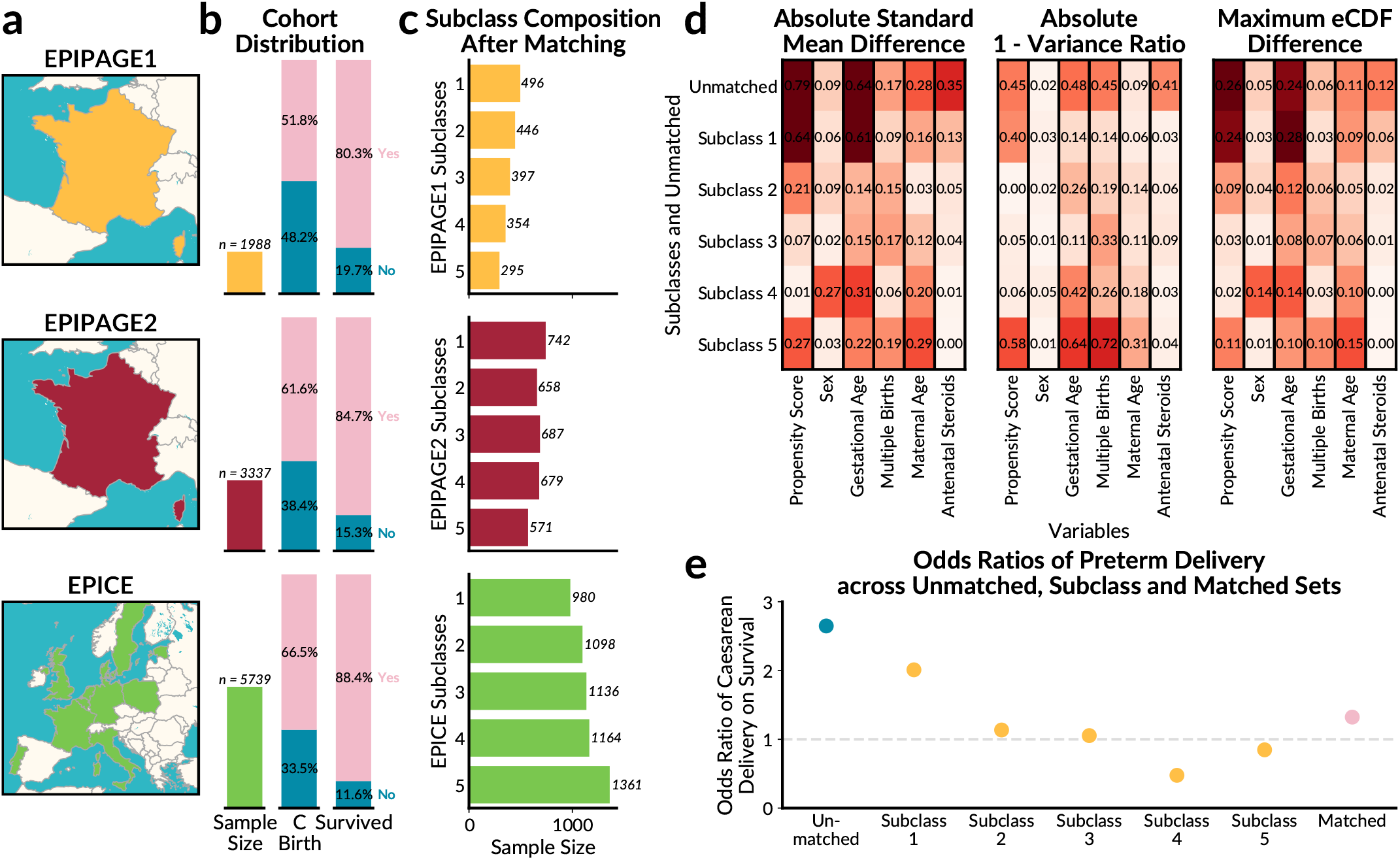
Secure Propensity Score Subclassification for Very Preterm Births in the RECAP Preterm platform. (a) Regions of the cohort studies available on the federated server node. (b) Overview of each respective cohort’s sample size, percentage of patients who underwent a Caesarean birth (C Birth) and number of patients who survived until discharge from hospital. (c) Number of samples chosen within each subclass of each cohort after secure propensity score subclass matching. (d) Heatmaps illustrating summary statistics of variables within each subclass as well as the unmatched dataset. (e) Odds Ratios for preterm delivery on survival outcomes which highlights a drop from the unmatched to the matched odds ratio after balancing of the cohorts.

Propensity score subclassification was used to balance maternal age, GA (weeks) at delivery, multiple birth status, child sex, and use of antenatal steroids amongst subjects with complete data. These variables were used to estimate the propensity of a Caesarean delivery and were used to split subjects into 5 subclasses (Figure 5c) as it produced sufficient covariate balance following the conventional use of five subclasses [Rosenbaum and Rubin, 1984]. After federated subclassification, we assessed the balance of Caesarean and vaginal birth populations across variables by computing the standard mean difference, variance ratio and maximum eCDF differences (Figure 5d). Each variable had generally lower summary values in the matched subclass set compared to the unmatched set, reflecting how federated subclassification improved the covariate balance.

After assessing the subclasses that offered sufficient balance in a secure manner, we used the generated weighted subclass samples to estimate the relative risk of Caesarean birth for child survival (Figure 5e). Again, we find that the naive analysis proposed an odds ratio of 2.65 of survival for Caesarean delivery. Yet, the odds ratio was substantially lower at 1.32 after federated subclassification, reflecting that the initial, unbalanced dataset had strong confounders for the mode of delivery. Our study here highlights how federated subclassification prevents deceptive results by facilitating an effective causal analysis pipeline whilst ensure data security.

## Discussion

In this work, we demonstrated how aspects of differential privacy and federated learning were used to develop the non-disclosive matching packages, *dsMatching* and *dsMatchingClient*, on the DataSHIELD platform. Our packages enable users to conduct and monitor covariate balancing across servers, thus allowing analysts to exploit the large number of samples from similar observational studies but whose access is limited due to privacy concerns. To this end, the proposed framework and the respective packages provide researchers with a variety of options for different balance specifications, diagnostic tools for evaluating covariate balance before and after matching, several plotting and visualisation aids, as well as estimation of average treatment effects and associated uncertainties. At the same time, data custodians also have the flexibility to choose the level of privacy they require.

In order to verify our methodology, we conducted a simulation study similar to well-known schemes employed by standard matching method research. We showed how the impact of differentially private-induced noise on the propensity score affects selected samples and average treatment. This unveils not only what trade-offs data custodians must make when specifying their privacy budget, but also what effect poorly estimated propensity scores generally have on estimands after matching. We then applied our package to two real-world datasets, both consisting of many consortia from similar studies. This allowed for evaluating covariate balance across consortia sites as well as estimating the treatment effect in a secure and reliable procedure, in contrast to site-restricted meta-analyses. As federated matching is performed on the pooled set of communicated propensity-score vectors, the matching algorithm itself is not site-specific. Thus, after successful communication and disclosure checks, increasing the number of sites does not directly affect the matching quality. However, site fragmentation can affect feasibility and runtime indirectly through extreme treatment imbalance, or suppression of outputs when site-level treatment groups fall below disclosure thresholds (Supplementary Table 1).

We note that the generated weighted samples after federated matching and subclassification can also be flexibly used in other federated tools. Therefore, our work should be viewed as complementary to other cross-site balancing methods such as federated inverse probability weighting, and federated doubly robust estimation [Xiong et al., 2023, Han et al., 2024, Yin et al., 2026]. Our methods focus on the covariate-balancing workflow itself: constructing matched or subclassified populations, retaining matched subsets locally, enabling privacy-preserving balance diagnostics, and providing treatment effect estimation on the resulting balanced population. Thus, federated matching and subclassification are most applicable when analysts require an interpretable matched cohort and direct assessment of covariate balance across multiple remote sites, while also mirroring established matching workflows such as those used in centralised tools like *MatchIt*. Future extensions could incorporate additional balancing strategies, such as coarsened exact matching or entropy balancing, particularly where analysts wish to reduce reliance on transmitting individual-level propensity-score representations.

Although our methodology has many strengths, there are some limitations. First, the proposed framework inherits the standard assumptions of propensity score methods. In particular, causal interpretation requires that all relevant confounders are observed and included in the propensity score model. This assumption cannot be verified empirically, and the presented applications should therefore be interpreted as adjusting for measured confounding rather than eliminating all possible confounding. Sensitivity analysis methods for unmeasured confounding such as Rosenbaum-type bounds are currently planned for future package versions. However, we highlight that sensitivity metrics like E-values can in principle be computed from risk-ratio estimates already produced by the package.

Second, while the randomised IDs and shuffling of propensity scores make recovery of data very difficult, if a malicious actor succeeds in circumventing these protections, servers with high privacy budgets will make recovery of data disturbed with minor noise possible, thereby increasing the risk of linkage or reconstruction attacks. To mitigate these risks, data custodians can restrict repeated calls to the server, review access logs, or require subclassification in settings where even shuffled, randomly-generated IDs are considered too sensitive.

Depending on the data custodian and study, a high privacy budget can be necessary in order to consistently recover the treatment effect and thus data leakage can also occur. Additionally, the values of the estimating function are aggregated across clusters per server in order to compute cluster-robust standard errors. In cases where there are many clustered samples not located in the same server, then relatively few values are aggregated and thus there may be a disclosure risk. Although extraction of private data from these estimating function values is not readily apparent, a malicious attacker may succeed as has been shown in other vulnerabilities [Huth et al., 2023]. Thus, data custodians can limit this functionality to comply with possible safety precautions.

## Conclusion

In summary, this work establishes privacy-preserving matching as a first-class component of federated causal analysis within DataSHIELD. By integrating established matching methodology with federated optimisation and differential privacy, dsMatching enables principled covariate balancing and treatment effect estimation without compromising data sovereignty. The results demonstrate that reliable recovery of causal estimands is achievable while retaining formal privacy guarantees, and that federated implementations can reproduce centralised inference. Beyond the specific applications presented, the proposed framework generalises to a broad class of observational and interventional studies in which data heterogeneity, regulatory constraints, and multicentre collaboration are intrinsic. As federated infrastructures become increasingly central to biomedical research, dsMatching provides a concrete, extensible foundation for secure causal inference across distributed cohorts.

## Author contributions statement

Roy Gusinow (R. G.) and Jan Hasenauer (J. H.) were responsible for conceptualizing the project and for the development of the methodology. R. G. handled the programming of software. Aline-Marie Florence (A. F.), Elisa Gentilotti (E. G.), Jade Ghosn (J. G.), Zaira R. Palacios-Baena (Z. R. P.), Adriana Tami (A. Tami), Alice Toschi (A. Toschi) and the ORCHESTRA Study Group were involved in patient recruitment and data collection for the ORCHESTRA dataset. Andrei Scott Morgan (A. S. M.) and Jennifer Zeitlin (J. Z.) were involved in data collection, data processing and analysis for the RECAP Preterm dataset. L. M. C., E. G., R. G., A. S. M, A. Tami and J. Z. managed and curated their respective datasets. L. M. C., R. G., J. H., Mihyeon Kim (M. K.) and A. S. M. were involved in writing the original draft and editing of the final version. L. M. C., R. G., J. H., and A. S. M. aided in the conceptualisation and visualisation of the figures.

## Acknowledgments

We thank all investigators, study coordinators, data managers, and members of the ORCHESTRA and RECAP Preterm study groups for their work in participant recruitment, data collection and data management. The full list of contributors is available in the Supplementary material.

This work was supported by the Deutsche Forschungsgemeinschaft (DFG, German Research Foundation) under Germany’s Excellence Strategy (EXC 2047—390685813, EXC 2151-390873048), and by the University of Bonn (via the Schlegel Professorship) [Jan Hasenauer]. The ORCHESTRA and RECAP Preterm project received funding from the European Union’s Horizon Europe Research and Innovation programme [grant numbers 101016167 and 733280 respectively]. The RECAP Preterm project has also been funded by the Horizon Europe Research and Innovation program [grant number 101156325: IMPROVE PRETERM project]. This work only reflects the views and opinions of the authors and not those of the European Commission or any of their bodies.

The authors used OpenAI’s GPT-5.5 to assist with language polishing, grammatical checks and editorial revision. All scientific content, analyses, interpretations, and final wording were completed by the authors, and where relevant, LLM-modified text was thoroughly reviewed and updated by the authors.

## Data Availability

The ORCHESTRA Long-Term Sequelae and RECAP Preterm data are not publicly available due to privacy and data governance constraints. Access may be granted upon reasonable request and subject to the respective data access procedures. The ORCHESTRA dataset may be requested at https://dataportal.orchestra-cohort.eu/dataaccess/ and data from the RECAP Preterm platform may be requested at https://platform.recap-preterm.eu.

