## Supplementary PDF for "Privacy-Preserving Matching for Federated Causal Inference in Multicentre Patient Cohorts"

This document provides supplementary information for the paper, "Privacy-Preserving Matching for Federated Causal Inference in Multicentre Patient Cohorts" and contains mathematical derivations, supplementary tables, as well as supplementary figures to provide added details presented in the paper. The authors used OpenAI's GPT-5.5 to assist with language polishing, grammatical checks and editorial revision. All scientific content, analyses, interpretations, and final wording were completed by the authors, and where relevant, LLM-modified text was thoroughly reviewed and updated by the authors.

#### 1 Study Group Membership

##### ORCHESTRA Study Group Members

The following additional representatives from each institution contributed to the development of the ORCHESTRA project.

###### INSERM

Amal Abrous, Delphine Bachelet, Marie Bartoli, Lila Bouadma, Minerva Cervantes-Gonzalez, Anissa Chair, Charlotte Charpentier, Sandrine Couffin-Cadiergues, Nathalie De Castro, Diane Descamps, Hang Doan, Céline Dorival, Xavier Duval, Hélène Esperou, François Goehringer, Ikram Houas, Isabelle Hoffmann, Salma Jaafoura, Simon Jamard, Cédric Laouénan, Nadhem Lahfej, Soizic Le Mestre, France Mentré, Christelle Paul, Aurélie Papadopoulos, Marion Schneider, Coralie Tardivon, Sarah Tubiana, Aurélie Wiedemann.

###### University of Verona (UNIVR)

Elena Addis, Maddalena Armellini, Anna Maria Azzini, Benedetta Barana, Lucia Bonato, Elena Carrara, Alessandro Castelli, Filippo Cioli Puviani, Michela Conti, Raffaella Cordioli, Carmine Cutone, Ruth Joanna Davis, Pasquale De Nardo, Miriam Emiliani, Alessio Esposito, Daniele Fasan, Giada Fasani, Giorgia Franchina, Jacopo Garlasco, Enrico Gibbin, Salvatore Hermes Dall'O', Chiara Konishi De Toffoli, Lorenza Lambertenghi, Federico Lattanzi, Andrea Leonardi, Francesco Luca, Gaia Maccarrone, Fulvia Mazzaferri, Massimo Mirandola, Matteo Morra, Alessandra Nazeri, Matilde Rocchi, Giulia Rosini, Chiara Perlini, Maria Diletta Pezzani, Laura Rovigo, Anna Giulia Salvadori, Andrea Sartori, Alessia Savoldi, Rebecca Scardellato, Marcella Sibani, Erica Sodano, Simona Sorbello, Lorenzo Tavernaro, Giorgia Tomassini, Alessandro Visentin, Stefania Vitali, Andrea Volpe, Chiara Zanchi, Gloria Mazzali, Giovanni Stabile, Gianluca Vantini, Riccardo Cecchetto, Davide Gibellini, Nicolò Cardobi, Debora Calì, Maria Paola Cecchini, Maddalena Marcanti, Anna Mason, Salvatore

Monaco, Marco Pattaro Zonta, Cinzia Perlini, Gianluigi Zanusso, Elda Righi, Mariana Nunes Pinho Guedes, Maria Mongardi, Concetta Sciammarella, Claudio Micheletto, Paolo Gisondi, Lidia Del Piccolo.

**University of Bologna (UNIBO)**

Natascia Caroccia, Cecilia Bonazzetti, Beatrice Tazza, Michela Di Chiara, Zeno Igor Adrien Pasquini, Domenico Marzolla, Giacomo Fornaro, Maddalena Giannella, Fabio Trapani, Lorenzo Marconi, Luciano Attard, Sara Tedeschi, Silvia Vituliano, Liliana Gabrielli, Tiziana Lazzarotto.

**Servicio Andaluz de Salud (SAS)**

Jesús Rodríguez-Baño, María Isabel Garcia Sánchez, Ana Belén Hidalgo Céspedes, Aurora Aleman Rodriguez, Lola Cubero Aranda, Paula Olivares Navarro, Sandra De la Rosa Riestra, José M. Bravo-Ferrero, Maria Giulia Caponcello.

**University Medical Center Groningen (UMCG)**

Gerolf de Boe, Bernardina T. F. van der Gun, María F. Vincenti-González, Alida C. M. Veloo, Daniele Pantano, Margriet van der Meer, Lilli Gard, Erley F. Lizarazo, Marjolein Knoester, Alex W. Friedrich, Hubert G. M. Niesters, Karin I. Wold.

**CINECA**

Salvatore Cataudella, Chiara Dellacasa, Elisa Rossi.

**University of Bonn**

Manuel Huth, Clemens Peiter.

#### RECAP Preterm Study Group

The following additional representatives from each institution contributed to the development of the RECAP Preterm platform and the collection of data for the EPIPAGE1, EPIPAGE2 and EPICE cohorts.

**Obstetric, Perinatal, Paediatric and Lifecourse Epidemiology (OPPaLE) team, Centre for Research in Epidemiology and Statistics (CRESS) UMR1153, INSERM, Paris, France**

Pierre-Yves Ancel, Valerie Benhammou, Majda Hamrit, Veronique Pierrat, Mariane Sentenac.

**EPIUnit– Instituto de Saúde Pública da Universidade do Porto, Porto, Portugal**

Henrique Barros.

**Clinical Care and Management Innovation Research Area, Bambino Gesù Children's Hospital, IRCCS, Rome, Italy**

Marina Cuttini.

**Department of Health Sciences, University of Leicester, Leicester, UK**

Elizabeth Draper, Samantha Johnson, Deborah Bamber, Helen E. Collins, Charlotte Powell, Bradley Manktelow.

**University of Antwerp, Antwerp, Belgium**

Patrick Van Reempts, Jo Lebeer.

**Children's Hospital, University Hospital, Philipps University Marburg, Marburg, Germany**

Rolf F Maier.

**Department of Neonatal Medicine, Karolinska University Hospital, Stockholm, Sweden**

Ulrika Ådén, Mikael Norman.

**University of Tartu, Tartu University Hospital, Tartu, Estonia**

Liis Toome, Heili Varendi.

**Department of Neonatology, Poznan University of Medical Sciences, Poznan, Poland**

Janusz Gadzinowski.

**Department of Neonatology, Hvidovre Hospital, Hvidovre, Denmark**

Pernille Pedersen.

**Department of Neonatology, Radboud University Medical Center, Nijmegen, the Netherlands**

Arno van Heijst.

**Department of Psychology, University of Warwick, Coventry, United Kingdom**

Nicole Baumann, Robert Eves, Marina Mendonça, Dieter Wolke.

**INESC TEC—Institute for Systems and Computer Engineering, Technology and Science, Porto, Portugal**

Gonçalo Campos Gonçalves, José Pedro Ornelas, João Correia Lopes, Artur Rocha.

**Global Foundation for Care of the Newborn Infant, Munich, Germany**

Ruth Kemper, Paloma Nosten, Nicole Thiele.

### 2 Supplementary Notes

This section serves as a supplement to the Methods section of the paper.

#### 2.1 Minimum Bin Threshold Check

Various server-side functions such as the subclassification (`ds.subclass`) and summary statistic (`ds.summary`) procedure perform the assignment or aggregations of samples along a specified variable, typically the treatment indicator or covariate level. In cases where the number of samples within a specified bin is too low, there exist a disclosure risk as samples can be identified easily. For example, the age of a given sample patient is equal to the mean age of the bin, if there exist only single sample in the bin (that is, the given sample patient themselves). Similar to how the base DataSHIELD functionalities operate (dsBase), after a server-side operation is performed, the output is checked against the environment variable `nfilter.subset` and if the number of samples within any bin is below such a threshold, the output is not returned to the central client. For a typical DataSHIELD/OPAL installation, the variable `nfilter.subset` equals 3 by default but can be changed by the custodian itself in order to provide appropriate flexibility for a given analysis.

#### 2.2 G-computation for server-side counterfactual predictions

Let $A_i \in \{0, 1\}$ denote the treatment indicator, where $A_i = 1$ corresponds to the intervention condition and $A_i = 0$ to the control condition. Let $X_i$ denote the observed covariates for individual $i$, and let $Y_i(1)$ and $Y_i(0)$ denote the corresponding potential outcomes. For a binary outcome, we model the probability of $Y_i$ using a generalised linear model,

$$Pr(Y_i \mid A_i, X_i) = g\left(\beta_0 + \beta_A A_i + X_i^\top \beta_X\right),$$

where $g(\cdot)$ is the inverse of the link function, chosen depending on the outcome (e.g. logit link for binary outcomes).

The coefficient vector, $\beta = (\beta_0, \beta_A, \beta_X)$ is estimated using the federated generalised linear model implemented in *ds.glm*. During model fitting, each server computes local contributions to the likelihood, score, or gradient using its own data, and only aggregated, non-disclosive quantities are communicated to the client. This yields a common set of global parameter estimates $\hat{\beta}$, without centralising individual-level outcomes, treatment indicators, or covariates.

After estimation, the fitted parameters $\hat{\beta}$ are used locally on each server to generate predicted potential outcomes for each individual under both treatment conditions. Specifically, for individual $i$, the server computes

$$\widehat{\mu}_i(1) = \widehat{\Pr}(Y_i = 1 \mid A_i = 1, X_i) = g\Big(\hat{\beta}_0 + \hat{\beta}_A + X_i^\top \hat{\beta}_X\Big),$$

and

$$\widehat{\mu}_i(0) = \widehat{\Pr}(Y_i = 1 \mid A_i = 0, X_i) = g\Big(\hat{\beta}_0 + X_i^\top \hat{\beta}_X\Big),$$

corresponding to the predicted probability of the outcome under treatment and control respectively.

These predictions are generated for every individual under both treatment conditions, irrespective of the treatment actually observed. The individual-level predicted values remain on the server on which the corresponding individual is stored. This is important because the fitted model parameters are shared across sites; consequently, disclosing sample-level predictions could reveal information about the local covariates.

We note that the aggregated quantity per server, $S$,

$$\widehat{\mu}^S = \sum_{i=1}^{N} \widehat{p}_i^S$$

is non-disclosive when the minimum bin threshold on the total number of samples, $N$ is met, and thus can be sent to the centralised server. Therefore, only aggregated, non-disclosive summaries of these predictions are returned to the client for subsequent causal effect estimation.

#### 2.3 Outcome Computation

The outcome of interest after matching can be considered to take the form, $f(A, X; \hat{\beta})$. Consider, for example, the average treatment effect (ATE), which is defined as

$$f(A, X; \hat{\beta}) = \widehat{\text{ATE}} = \mathbb{E}[Y_i(1) - Y_i(0) \mid A, X; \hat{\beta}].$$

Let $C$ denote the target population over which the causal effect is averaged. For the ATE, $C = \{1, \ldots, n\}$. Let $w_i^C$ be the matching or subclassification weight for an individual $i$ in the population $C$, and so the total weight is $W_C = \sum_{i \in C} w_i^C$.

Using model-based standardization after matching, the estimated mean potential outcome under treatment level $d \in \{0, 1\}$ is

$$\widehat{\mu}_d^C = \frac{\sum_{i \in C} w_i^C \widehat{p}_i(d)}{\sum_{i \in C} w_i^C} = \frac{1}{W_C} \sum_{i \in C} w_i^C \widehat{p}_i(d).$$

Therefore, the ATE is estimated as

$$\widehat{\text{ATE}} = \widehat{\mu}_1^C - \widehat{\mu}_0^C = \frac{1}{W_C} \sum_{i \in C} w_i^C \left[\widehat{p}_i(1) - \widehat{p}_i(0)\right].$$

Similarly, the average treatment effect among the untreated estimate, $\widehat{\text{ATU}}$, and the average treatment effect among the treated, $\widehat{\text{ATT}}$, can be expressed as

$$\widehat{\text{ATU}} = \widehat{\mu}_1^{C_0} - \widehat{\mu}_0^{C_0}, \qquad \widehat{\text{ATT}} = \widehat{\mu}_1^{C_1} - \widehat{\mu}_0^{C_1},$$

where $C_1 = \{i : A_i = 1\}$ for the ATT and $C_0 = \{i : A_i = 0\}$ for the ATU, using the corresponding matching or subclassification weights for the chosen estimand.

Here, $\widehat{\mu}_1^C$ is the estimated mean outcome risk if all individuals in the target population $C$ were assigned to treatment, and $\widehat{\mu}_0^C$ is the estimated mean outcome risk if all individuals in $C$ were assigned to control.

In the federated setting, individual-level predicted potential outcomes remain on the remote servers. Let $s = 1, \ldots, S$ index servers and let $h = 1, \ldots, H$ index matched groups or subclasses. Each server computes only weighted aggregate quantities,

$$U_{s,d}^C = \sum_{h=1}^{H} \sum_{i \in C_{s,h}} w_i^C \widehat{p}_i(d), \qquad V_s^C = \sum_{h=1}^{H} \sum_{i \in C_{s,h}} w_i^C, \qquad \Longrightarrow \text{ computed at server } s.$$

for treatment indicator, $d \in \{0, 1\}$, where $C_{s,h}$ is the subset of individuals in target population $C$ stored on server $s$ and belonging to matched group or subclass $h$. The client then computes

$$\widehat{\mu}_d^C = \frac{\sum_{s=1}^{S} U_{s,d}^C}{\sum_{s=1}^{S} V_s^C}, \qquad \widehat{\tau}^C = \widehat{\mu}_1^C - \widehat{\mu}_0^C, \qquad \Longrightarrow \text{ computed at central client.}$$

In addition, the log risk ratio, $\log \widehat{RR}^C$, and log odds ratio, $\log \widehat{OR}^C$, in target population $C$ are

$$\log \widehat{RR}^C = \log \frac{\widehat{\mu}_1^C}{\widehat{\mu}_0^C} \quad \text{and} \quad \log \widehat{OR}^C = \log \frac{\widehat{\mu}_1^C/(1 - \widehat{\mu}_1^C)}{\widehat{\mu}_0^C/(1 - \widehat{\mu}_0^C)}.$$

##### 2.4 Uncertainty Quantification of the Outcome

Calculation of the cluster-robust standard errors for the ATE and other outcomes are necessary as estimates are correlated after matching. Here, the Delta method is utilised, in which we wish to estimate the variance. Using a first-order Taylor approximation, the variance of the outcome function, for example the ATE, is,

$$\text{Var}\left[f\left(X; \hat{\beta}\right)\right] \approx J^T V J,$$

where $J = \nabla_{\hat{\beta}} f\left(X; \hat{\beta}\right)$ is the Jacobian of the ATE outcome function with respect to the $\hat{\beta}$ and $V$ is the cluster-robust variance covariance matrix of the fitted parameter vector $\hat{\beta}$.

###### 2.4.1 Jacobian Estimation through Multiple Federated Nodes

$J = \nabla_{\hat{\beta}} f$ is simply the gradient of the outcome function with respect to the estimated parameters $\hat{\beta}$. We can be numerically derive the $k$-th component with a given step size $\varepsilon$,

$$J_k \approx \frac{f(A, X; \hat{\beta}_k + \varepsilon e_k) - f(A, X; \hat{\beta}_k)}{\varepsilon},$$

where $e_k$ is the $k-$th unit vector. We reiterate here that since the expression for the outcome function in securely aggregated per server, an expression of the gradient is similarly non-disclosive when shared at a centralised site.

##### 2.4.2 Heteroskedasticity-consistent Covariance Estimation with the Sandwich Estimator

In typical research areas which utilise matching, such as medicine, observations may not have the same uncertainty associated across the study. In the context where multiple data centers may house data collected through different methodologies and procedures, it is important that the accuracy of observation between datasets stored in different remote centers is accounted for when conducting federated analyses. Therefore, a heteroskedastic-consistent variance-covariance matrix is estimated using the sandwich estimator [1]. The variance-covariance matrix $V$ can be derived with the sandwich estimator,

$$V = \hat{A}^{-1}\hat{B}\hat{A}^{-1},$$

where the "bread" estimate $\hat{A}$ is based on the empirical version of the inverse Hessian of the objective function, $\Psi(y, x, \beta)$ in the case of a GLM. That is,

$$\hat{A}(\hat{\beta}) = \mathbb{E}\left[-\frac{\partial\psi(Y, X, \hat{\beta})}{\partial\hat{\beta}}\right]$$

Thus, $\hat{A}\left(\hat{\beta}\right)$ is equivalent to the covariance of the regression on $X$, after model fitting of $f(A, X; \hat{\beta})$.

We may use the Eicker-Huber-White estimator to calculate the meat matrix $\hat{B} = Var[\psi]$ as the outer product of the estimating functions. In the case of a least square problem, the estimation function $\psi(Y, X; \hat{\beta})$ is equal to the derivative of the objective function $\Psi(Y, X; \hat{\beta})$, such that $\hat{\beta}$ is the argument that gives the minimum value. In the federated setting, individual-level outcomes, covariates, and predicted potential outcomes remain on the remote servers. Let $s = 1, \ldots, S$ index the servers and $h = 1, \ldots, H$ indicate the subclasses or matched groups obtained after the balancing procedure, so the meat matrix is computed as,

$$\hat{B} = \frac{1}{N}\sum_{s=1}^{S}\sum_{h=1}^{H}\left(\sum_{i=1}^{n_{s,h}}\psi\left(Y_{s,h,i}, X_{s,h,i}; \hat{\beta}\right)\right)\left(\sum_{i=1}^{n_{s,h}}\psi\left(Y_{s,h,i}, X_{s,h,i}; \hat{\beta}\right)\right)^T.$$

The difficulty in providing privacy guarantees in this formulation is that the summation of estimating functions before the outer product involves taking samples within the same cluster, $h$ within the same servers, $S$. For 1 to 1 matching, where $H$ is large, it is likely that matched paired exist across servers, and thus the estimating function values, $X_{s,h,i}(Y_{s,h,i} - \hat{\beta}X_{s,h,i})$ have to be transmitted to the central server before the outer product computation. Although the sample's covariate vector, $X_{s,h,i}$ and the residual $Y_{s,h,i} - \hat{\beta}X_{s,h,i}$ are not known to the central server, and thus cannot be recovered directly from the estimating function values, the operation requires returning a matrix the dimensions of the design matrix, $X$. However, in subclassification matching, $H$ is small, with stratum divide by global propensity score quantiles, and so, we are likely to aggregate stratum groups before communication to the central site, thus preserving data security.

Overall, in the implementation provided, users are given the option to decide on which level of data security they would like to employ when requesting uncertainty estimates through the average treatment effect, using a choice of 1 to 1 or subclassification matching on the propensity scores.

#### 2.5 Federated propensity-score matching objective

Federated matching follows the same distance-based logic as the standard centralised propensity-score matching [2], but replaces the unperturbed propensity scores, $\pi_i$ with differentially private propensity-score representations, $\tilde{\pi}_i$. Let $i$ index individuals within a site, so each individual has a treatment indicator $A_i \in \{0, 1\}$ and an estimated propensity score $\pi_i$, computed locally at the remote site.

Before matching, each site constructs a temporary randomised identifier $r_i$, perturbs the propensity score to obtain $\tilde{\pi}_i$, shuffles the local records, and transmits only $(r_i, D_i, \tilde{\pi}_i)$ to the central client. The original covariates, outcomes, local record identifiers, and unperturbed propensity scores remain on the remote servers.

The propensity-score distance between treated unit, $j \in \mathcal{T}$ and control unit, $k \in \mathcal{C}$ is defined as

$$d_{jk} = \left|\tilde{\pi}_j - \tilde{\pi}_k\right|,$$

noting that the treated group, $\mathcal{T}$ denotes the set of all samples where all $D_j = 1$, and conversely the control set, $\mathcal{C}$ has samples, $D_k = 0$. Restrictions, such as discarding observations outside the region of common support, are applied before constructing the final set of admissible pairs.

For one-to-one nearest-neighbour matching without replacement, treated units are processed according to descending ordering, and each treated unit $i$ is progressively paired off with a corresponding the admissible control unit with the smallest value of $d_{jk}$. This can be written as

$$m(j) = \arg\min_{k \in \mathcal{C}_j} d_{jk},$$

where $\mathcal{C}_i$ is the set of controls currently still available for treated unit $i$. If matching is performed with replacement, previously selected controls remain available for later treated units, and so the same control unit may be reused for multiple treated units. If matching is performed without replacement, selected controls are removed from the available control set, $\mathcal{C}_i$.

For optimal pair matching, the client instead selects binary assignment variables $a_{jk} \in \{0, 1\}$ to minimise the total matched distance,

$$\min_{\{a_{jk}\}} \sum_{j \in \mathcal{T}} \sum_{k \in \mathcal{C}} a_{jk} d_{jk},$$

subject to the selected matching constraints, such as the matching ratio, discarding choice, and whether controls may be reused.

Cross-site matches are permitted here because matching is performed on the pooled set of communicated temporary identifiers and perturbed propensity scores. Therefore, a treated individual from one site may be matched to a control individual from another site. After the client identifies the selected matched records, only the corresponding temporary identifiers are returned to the relevant remote servers. Each server then constructs the matched-subset indicator locally. Subsequent balance diagnostics and treatment-effect estimation are performed through federated aggregation, so individual-level outcomes and covariates are not transmitted to the client.

#### 2.6 Pseudocode

This section details the pseudocode of functions implemented in the DataSHIELD R package, detailing where aggregations of sample information is made and how these computations are summarised in the central client.

To ensure correctness of the methodology under a variety of matching specifications and outcome effects, the testthat configuration of our R DataSHIELD package compares the centralised and federated implementations using their corresponding the results generated [3]. The test cases use the ”lalonde” dataset [4] typically used for demonstrating calculations of causal effect estimation and shown in the MatchIt package [2]. In order to appropriately simulate the capabilities of our package on remote servers, we employ DSLite, a serverless implementation used to mimic the behaviour of the DataSHIELD installation which would be on a remote site. In cases where noisy propensity scores are used, the privacy budget has been set to $10^5$. For these comparisons, the centralised and federated results are expect to agree up to the $10^{-3}$ tolerance specified.

##### ds.matchit

**Algorithm 1 ds.matchit**: Federated propensity-score matching (server scores, client matching)

**Input:** Formula $f$, server table name `data`, output name `newobj`, MatchIt options . . .
**Output:** Matched dataset assigned on each server as `newobj`; vector `subclass` returned to client

**Add stable identifier on each server:**
  `assign_IDDS(data, id_name="ID")`

**Estimate propensity model (federated):**
  $\hat{\beta}$ ← `ds.glm(f, data, family="binomial")`

**Compute propensity scores on servers (using pooled coefficients):**
  `ds.genProp(f, coefficients=`$\hat{\beta}$`, data, newobj="distance")`
  `ds.cbind(data, "distance", newobj=data)` ▷ append score to server table

**Get differentially-private scores per server and pool them:**
**for** each server $s \in \{1, \ldots, n\}$ **do**
    `noisy_pool[`$s$`]` ← `matchitDS(f, data, "distance", "ID")` ▷ returns ID + noisy distance on server $s$
**end for**
  `prop_pool` ← `pool_servers(noisy_pool)` ▷ join into single dataframe

**Client-side matching (MatchIt):**
  `fit` ← `MatchIt::matchit(f, prop_pool, ...)`
  `matched` ← `MatchIt::match.data(fit)` ▷ contains ID, weights, subclass, distance

**Push match results back to servers:**
  `matchitDS2(data, matched$ID, matched$distance, matched$weights, matched$subclass)`

#### ds.matchit_subclass

**Algorithm 2 ds.matchit_subclass**: Federated propensity-score subclassification (server scores, pooled cutpoints)

**Input:** Formula $f$, server table name `data`, number of subclasses $K$, estimand $\in$ {ATE, ATT, ATU}, output name `newobj`

**Output:** Subclassified dataset assigned on each server as `newobj` (with weights); client returns balance summary, sample sizes, cutpoints, and subclass labels

**Estimate propensity model (federated):**
  $\hat{\beta} \leftarrow$ `ds.glm(f, data, family="binomial")`

**Compute propensity scores on servers (using pooled coefficients):**
  `ds.genProp(f, coefficients=`$\hat{\beta}$`, data, newobj="distance")`
  `ds.cbind(data, "distance", newobj=data)` ▷ append score to server table

**Get quantile cutpoints (federated):**
**if** `estimand = "ATT"` **then**
    `domain` $\leftarrow$ subset(`data` to treated units)
**else if** `estimand = "ATU"` **then**
    `domain` $\leftarrow$ subset(`data` to control units)
**else**
    `domain` $\leftarrow$ `data` ▷ treated + control units
**end if**
  `quantiles` $\leftarrow$ `ds.quantile(domain, K + 1)` ▷ global cutpoints

**Assign subclasses on each server using pooled cutpoints:**
  `matchit_subclassDS(data, distance="data$distance", quantiles)` $\rightarrow$ `newobj`

**Summarise balance and subclass sample sizes (federated):**
  `summary` $\leftarrow$ `ds.match_summary_subclass(data, newobj)` ▷ per subclass: control, treated, total

**Compute subclass weights (depends on estimand):**
**if** `estimand = "ATT"` **then**
    `weights_list[k]` $\leftarrow \frac{\#T_k}{\#C_k}$ ▷ controls reweighted within subclass $k$
**else if** `estimand = "ATU"` **then**
    `weights_list[k]` $\leftarrow \frac{\#C_k}{\#T_k}$ ▷ treated reweighted within subclass $k$
**else**
    `weights_list[T,k]` $\leftarrow \frac{\#N_k}{\#T_k}$; `weights_list[C,k]` $\leftarrow \frac{\#N_k}{\#C_k}$ ▷ both groups reweighted within subclass $k$
**end if**

**Assign weights back onto servers:**
  `weights` $\leftarrow$ `assign_weights_subclassDS(newobj, weights_list, estimand)`

**Return outputs to client:**
  `summary$quantiles` $\leftarrow$ `quantiles`
  `summary$subclass` $\leftarrow$ vector of subclass labels for all matched rows
**return** `summary`

#### Federated Treatment Effect

**Algorithm 3** Federated Average Treatment Effect (ATE) estimation after matching / subclassification

**Input:** outcome model $f$ for predictions using parameters $\beta$; servers $s = 1, \ldots, S$ holding $(Y, X, A)$; matched groups / subclasses $h = 1, \ldots, H$; target estimand ATE

**Output:** Global $\widehat{ATE}$ computed on client from server-level aggregates

**G-computation on each server (counterfactual predictions):**
  Create two counterfactual copies of the matched data:
    `data_1`: set treatment $A \leftarrow 1$ for all rows
    `data_0`: set treatment $A \leftarrow 0$ for all rows
  Compute predicted outcomes under each treatment level:
    $\hat{y}_{d=1} \leftarrow \hat{y}(X, A{=}1; \beta)$ using `data_1`
    $\hat{y}_{d=0} \leftarrow \hat{y}(X, A{=}0; \beta)$ using `data_0` ▷ server-side prediction

**Within-server, within-class aggregated summaries:**
  For each server $s$ and class $h$, compute weighted summaries over the target population $C$:
    $W^C_{s,h} \leftarrow \sum_{i \in C_{s,h}} w^C_i$ ▷ sum of matching/subclassification weights
    $S^C_{s,h,1} \leftarrow \sum_{i \in C_{s,h}} w^C_i \hat{y}_{s,h,i}(1)$
    $S^C_{s,h,0} \leftarrow \sum_{i \in C_{s,h}} w^C_i \hat{y}_{s,h,i}(0)$ ▷ weighted sums of predicted potential outcomes

**Client-side aggregation and ATE:**
  $S^C_1 \leftarrow \sum_{s=1}^{S} \sum_{h=1}^{H} S^C_{s,h,1}$ and $S^C_0 \leftarrow \sum_{s=1}^{S} \sum_{h=1}^{H} S^C_{s,h,0}$ ▷ weighted sums across servers and classes
  $W_C \leftarrow \sum_{s=1}^{S} \sum_{h=1}^{H} W^C_{s,h}$ ▷ total target-population weight
  $\widehat{\mu}^C_1 \leftarrow S^C_1 / W_C$ and $\widehat{\mu}^C_0 \leftarrow S^C_0 / W_C$
  $\widehat{ATE} \leftarrow \widehat{\mu}^C_1 - \widehat{\mu}^C_0$

**return** $\widehat{ATE}$

#### Federated Treatment Effect Uncertainty

**Algorithm 4** Cluster-robust uncertainty for federated ATE via Delta method and sandwich estimator

**Input:** Matched/subclassified data inducing clusters $h$; fitted global parameters $\hat{\beta}$; step size $\varepsilon$; cluster definition (pairs for 1:1 matching or strata for subclassification)

**Output:** Cluster-robust standard error for $\widehat{ATE}$

**Delta method structure:**
 $\widehat{\mathrm{Var}}(\widehat{ATE}) \approx J^{\mathsf{T}} V J$
 where $J = \nabla_{\beta}\, ATE(X; \beta)\big|_{\beta=\hat{\beta}}$ and $V$ is cluster-robust $\mathrm{Var}(\hat{\beta})$

**Numerical Jacobian (federated forward difference):**
**for** each parameter component $k$ of $\beta$ **do**
  `client picks perturbation:` $\beta^{(+)} \leftarrow \hat{\beta}$ with $\beta_k^{(+)} \leftarrow \hat{\beta}_k + \varepsilon$
  `client requests federated ATE at` $\hat{\beta}$ and at $\beta^{(+)}$ (Algorithm 3)
  $J_k \leftarrow \dfrac{\widehat{ATE}(X; \beta^{(+)}) - \widehat{ATE}(X; \hat{\beta})}{\varepsilon}$
**end for**

**Sandwich covariance with clustering:**
 $V \leftarrow \hat{A}^{-1}\, \hat{B}\, \hat{A}^{-1}$
 $\hat{A}$ obtained from model information (`ds.glm`) in aggregated fashion

**Meat aggregation by clusters:**
**for** each server $s \in \{1, \ldots, S\}$ **do**
 **for** each cluster $h$ on server $s$ **do**
  Compute cluster score/estimating-function sum:
   $g_{s,h} \leftarrow \sum_{i \in (s,h)} \psi(y_{s,h,i}, x_{s,h,i}, \hat{\beta})$
  `send_to_client:` $g_{s,h}$
 **end for**
**end for**
 `client forms:` $\hat{B} \leftarrow \dfrac{1}{N} \sum_{s=1}^{S} \sum_{h=1}^{H} g_{s,h}\, g_{s,h}^{\mathsf{T}}$

**Variance and standard error:**
 $\widehat{\mathrm{Var}}(\widehat{ATE}) \leftarrow J^{\mathsf{T}} V J$
 $SE(\widehat{ATE}) \leftarrow \sqrt{\widehat{\mathrm{Var}}(\widehat{ATE})}$

**return** $SE(\widehat{ATE})$

### 3 Supplementary Figures

This section supplies additional figures which support the primary text.

#### 3.1 Central and Federated Comparisons

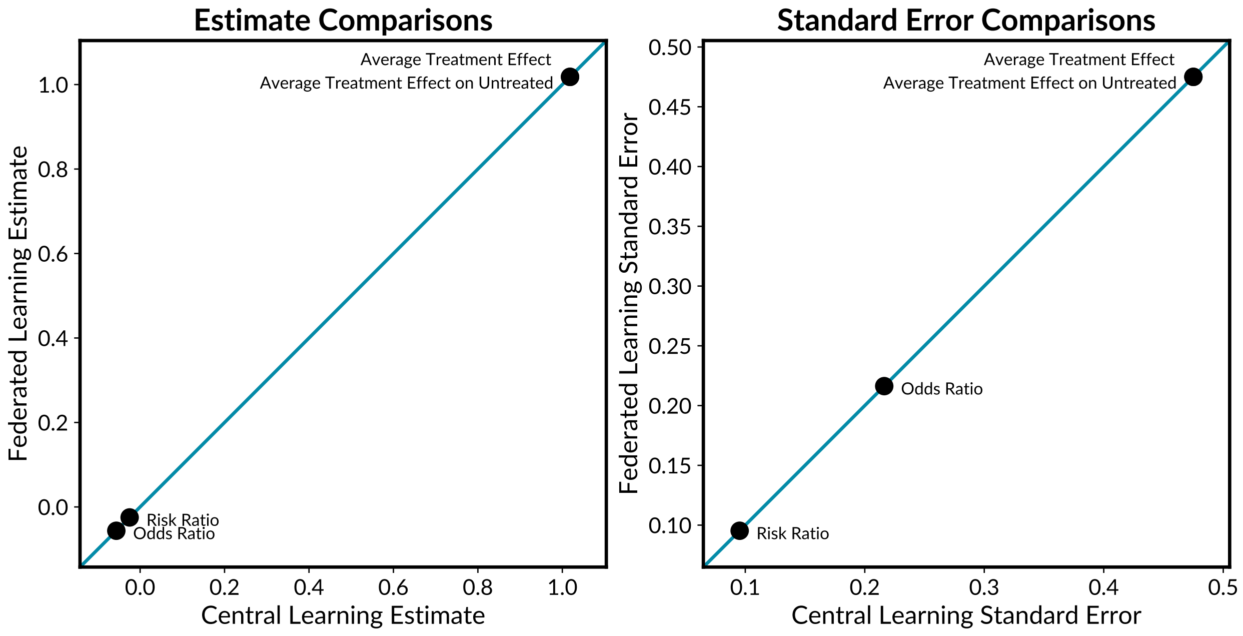


Supplementary Figure 1: **Central and Federated Comparisons for Various Outcomes.** The Average Treatment Effect (ATE), Average Treatment Effect on Untreated (ATU), Risk Ratio (RR) and Odds Ratio is estimated on the simulated dataset after nearest neighbour matching for both the central and federated estimate. Both results yield the same values up to numerical error.

### 4 Supplementary Tables

| Size | Server Count | GLM Time (s) | Matching Time (s) | Outcome Time (s) | Total Time (s) |
|---|---|---|---|---|---|
| 1,000 | 3 | 1.24 | 1.57 | 6.71 | 9.52 |
| 1,000 | 5 | 2.11 | 2.53 | 11.27 | 15.91 |
| 1,000 | 10 | 3.87 | 4.90 | 21.52 | 30.30 |
| 10,000 | 3 | 1.24 | 2.00 | 7.04 | 10.28 |
| 10,000 | 5 | 2.25 | 3.30 | 12.72 | 18.27 |
| 10,000 | 10 | 4.05 | 5.95 | 23.47 | 33.47 |
| 50,000 | 3 | 2.00 | 6.31 | 8.21 | 16.51 |
| 50,000 | 5 | 2.51 | 6.86 | 13.81 | 23.19 |
| 50,000 | 10 | 4.09 | 10.36 | 24.37 | 38.83 |

Supplementary Table 1: **Runtime performance averaged across 10 runs using remote DataSHIELD servers.** For a simulated dataset based on Figure 3 of the main manuscript, nearest-neighbour federated matching was performed and the corresponding ATT was estimated. In each configuration, the total sample size was partitioned equally across the specified number of servers, and elapsed time in seconds was recorded over 10 independent runs. Reported timings summarise generalised linear model fitting and propensity score generation (GLM), transmission and matching operations (Matching), outcome estimation (Outcome), and overall runtime (Total). Benchmarks were conducted on an Ubuntu 24.04.2 LTS operating system with an Intel Core i7-1185G7 processor; at the time of testing, the network connection measured 828.08 Mbps download, 256.70 Mbps upload, and 11.17 ms latency. Runtime increased with both the number of servers and the total sample size, with outcome estimation generally contributing the largest share of total computation time.
